# Japanese Project for Telepsychiatry Evaluation during COVID-19: Treatment Comparison Trial (J-PROTECT): Rationale, design, and methodology

**DOI:** 10.1101/2021.06.23.21259366

**Authors:** Taishiro Kishimoto, Shotaro Kinoshita, Shogyoku Bun, Yasunori Sato, Momoko Kitazawa, Toshiaki Kikuchi, Mitsuhiro Sado, Akihiro Takamiya, Masaru Mimura, Takashi Nakamae, Yoshinari Abe, Tetsufumi Kanazawa, Yasuo Kawabata, Hiroaki Tomita, Koichi Abe, Akitoyo Hishimoto, Takeshi Asami, Akira Suda, Yoshinori Watanabe, Toru Amagai, Kei Sakuma, Hisashi Kida, Michitaka Funayama, Hiroshi Kimura, Aiko Sato, Shuichiro Fujiwara, Kiichiro Nagao, Naoya Sugiyama, Maki Takamiya, Hideyuki Kodama, Takaharu Azekawa, on behalf of the J-PROTECT collaborators

## Abstract

**Introduction:** The COVID-19 pandemic has had a profound impact on the mental health of people around the world. Anxiety related to infection, stress and stigma caused by the forced changes in daily life have reportedly increased the incidence and symptoms of depression, anxiety disorder and obsessive-compulsive disorder. Under such circumstances, telepsychiatry is gaining importance and attracting a great deal of attention. However, few large pragmatic clinical trials on the use of telepsychiatry targeting multiple psychiatric disorders have been conducted to date.

**Methods:** The targeted study cohort will consist of adults (>18 years) who meet the DSM-5 diagnostic criteria for either (1) depressive disorders, (2) anxiety disorders, or (3) obsessive-compulsive and related disorders. Patients will be assigned in a 1:1 ratio to either a “telepsychiatry group” (at least 50% of visits to be conducted using telemedicine, with at least one face-to-face treatment [FTF] every six months) or an “FTF group” (all visits to be conducted FTF, with no telemedicine). Both groups will receive the usual treatment covered by public medical insurance. The study will utilize a master protocol design in that there will be primary and secondary outcomes for the entire group regardless of diagnosis, as well as the outcomes for each individual disorder group.

**Discussion:** This study will be a non-inferiority trial to test that the treatment effect of telepsychiatry is not inferior to that of FTF alone. This study will provide useful insights into the effect of the COVID-19 pandemic on the practice of psychiatry.

**Trial Registration:** jRCT1030210037, Japan Registry of Clinical Trials (jRCT)

## 1. INTRODUCTION

The 2019 novel coronavirus disease (COVID-19) pandemic, which is raging worldwide, has had significant direct and indirect impacts on mental health. The incidence of symptoms, such as anxiety, depression, and obsessive–compulsive disorder (OCD), in the general population is reportedly higher than during pre-pandemic times as a result of the effects of lockdowns and other restrictions on going out, sudden unemployment, and frequent exposure to related topics through news associated with the COVID-19 pandemic [1–2]. People infected with COVID-19 also reportedly have a higher risk of developing psychiatric disorders, and people with a history of psychiatric disorders have a higher diagnosis rate of COVID-19 infection [3]. This situation suggests that psychiatric care is becoming increasingly important under the COVID-19 pandemic.

However, during the COVID-19 pandemic, patients may refrain from visiting hospitals because of fears of infection, and hospitals may stop performing outpatient consultations. According to a survey conducted by WHO in 130 countries, 93% of the countries surveyed reported obstacles to providing psychiatric and mental health services [4]. Under these circumstances, it is important to find ways to continue the provision of psychiatric care. In past epidemics of infectious diseases, telemedicine has been used to prevent infection among healthcare workers by using the telephone and video calls to conduct consultations [5]. During the recent pandemic, telemedicine has attracted attention as a way of continuing medical care while preventing infection, and its use has rapidly expanded due to deregulation in many countries [6].

In psychiatry, outpatient care is mainly conducted in the form of in-person conversations; therefore, doctor-to-patient telemedicine, in which patients are treated remotely via videophone, is easy to apply in this field. For this reason, telemedicine was popular in the field of psychiatry even before the COVID-19 pandemic [7]. There have also been many studies comparing the effectiveness of telepsychiatry with face-to-face (FTF) treatment, and telepsychiatry can reportedly provide equivalent or better treatment effects, patient satisfaction, and improved medication adherence, compared with FTF treatment [8–10]. However, most of the existing studies have focused on only a single disorder, such as depression or PTSD, or on experimental trials that differ from real-world clinical practice, and few large pragmatic trials examining multiple psychiatric disorders have been reported worldwide.

As COVID-19 is expected to take some time to fully resolve, verifying whether telemedicine is as effective as FTF treatment in real-world clinical practice for the treatment of depression, anxiety, and OCD in existing clinical settings is of great importance, particularly since increases in the diagnosis or exacerbation of these conditions have been reported in response to the circumstances created by the COVID-19 pandemic.

## 2. Method

### 2.1. Design Overview

Patients will be assigned in a 1:1 ratio to either a “telepsychiatry group” (at least 50% of visits to be conducted by telemedicine, with at least one FTF every six months) or an “FTF group” (all visits to be FTF, with no telemedicine). Both groups will receive the usual treatment covered by public medical insurance. The interval between visits will be at the discretion of the psychiatrist in charge. This study will have a master protocol with primary and secondary outcomes for the entire group as well as outcomes for each individual disorder group.

The main objective of this study will be to show that the telepsychiatry group is non-inferior to the FTF group after 6 months of practice in patients with depressive disorder, anxiety disorder, or OCD and related disorders.

### 2.2. Participants

This study will be a multi-site, prospective randomized controlled trial. Participants will be recruited at 17 medical institutions that provide psychiatric services in 11 different prefectures in Japan. Patient recruitment will be conducted at the following locations and medical institutions: Tokyo (Keio University Hospital, Himorogi Psychiatric Institute); Kanagawa (Yokohama City University Hospital, Shioiri Mental Clinic, Amagai Mental Clinic, Kanazawabunko Yell Clinic); Fukushima (Asaka Hospital); Miyagi (Tohoku University Hospital); Kyoto (University Hospital Kyoto Prefectural University of Medicine); Osaka (Osaka Medical and Pharmaceutical University Hospital, Neyagawa Sanatorium); Tochigi (Ashikaga Red Cross Hospital); Yamagata (Sato Hospital); Miyazaki (Takamiya Hospital); Shizuoka (Numazu Chuo Hospital); and Chiba (International University of Health and Welfare University Narita Hospital, Gakuji-kai Kimura Hospital).

The inclusion criteria for participants will be as follows: 1) patients who meet the Diagnostic and Statistical Manual of Mental Disorders, Fifth Edition (DSM-5) criteria for depressive disorders, anxiety disorders, or OCD and related disorders and are outpatients at a participating medical institution; 2) patients who are 18 years old or older at the time of obtaining consent; 3) patients who need continuous treatment for the next 6 months or more (at the discretion of the attending physician); 4) patients who have a smart phone or PC and have access to video calling over the Internet (even if it is only available with family support); 5) patients whose psychiatric conditions are stable enough to receive telepsychiatry by clinical judgement of the attending physician; 6) patients whose psychiatric conditions are stable enough to have sufficient capacity provide consent by clinical judgement of the attending physician; and 7) patients who have provided written consent to participate in the study. For patients who are minors (less than 20 years of age), written consent must be obtained from the patient and his/her guardian. Exclusion criteria include: 1) patients who are likely to require unscheduled or urgent visits to the hospital in addition to regular visits because of emergent suicidal ideation, anxiety or agitation; and 2) patients who have difficulty managing an emergency visit by themselves when their psychiatric conditions deteriorate (e.g., the hospital is located far away). Researchers will obtain written informed consent from all the participants. The participants will be able to leave the study at any time.

### 2.3. Randomization

Patients who have given their consent to participate in this study will be randomly assigned in a 1:1 ratio to the “Telepsychiatry group” or the “FTF group” for treatment during the study period. In this study, to avoid inter-institutional differences and biases in the types of disorders among the groups, randomization process is carried out by a blinded, independent third party using a modified minimization method with a biased-coin assignment balanced for age group (60 years old or older, or younger), gender (male or female), target disorder, and participating institution. In addition, the allocation results will not be disclosed to the central evaluator to minimize bias.

### 2.4. Assessment Schedule

The assessment schedule is presented in Table 1. After randomization, participants will be assessed with the SF36MCS at baseline and at weeks 12 and 24. The duration of each patient’s participation in the study is estimated to be approximately 6 to 7 months, including the allowance period for the prescribed visits.

**Table 1.**
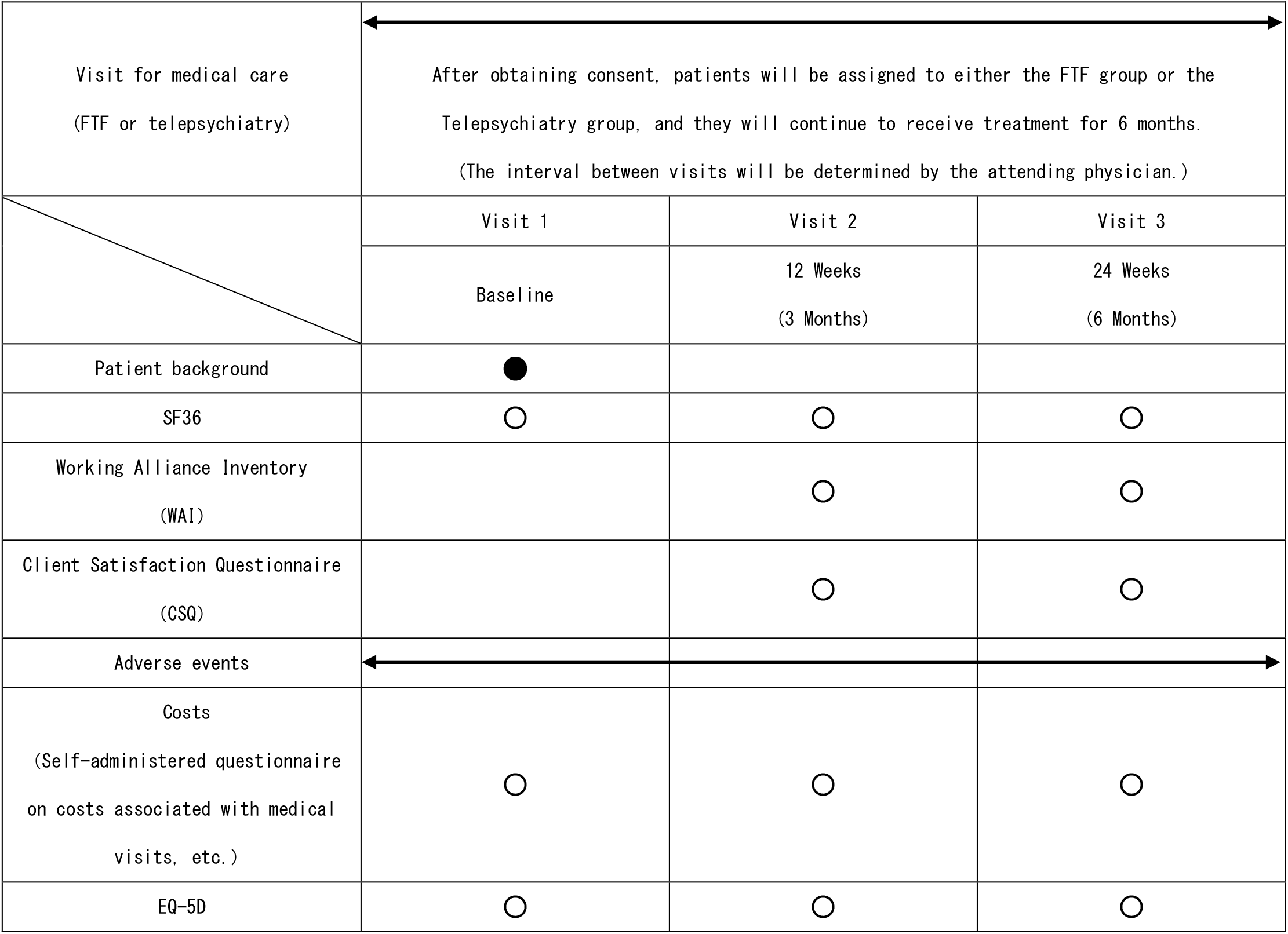
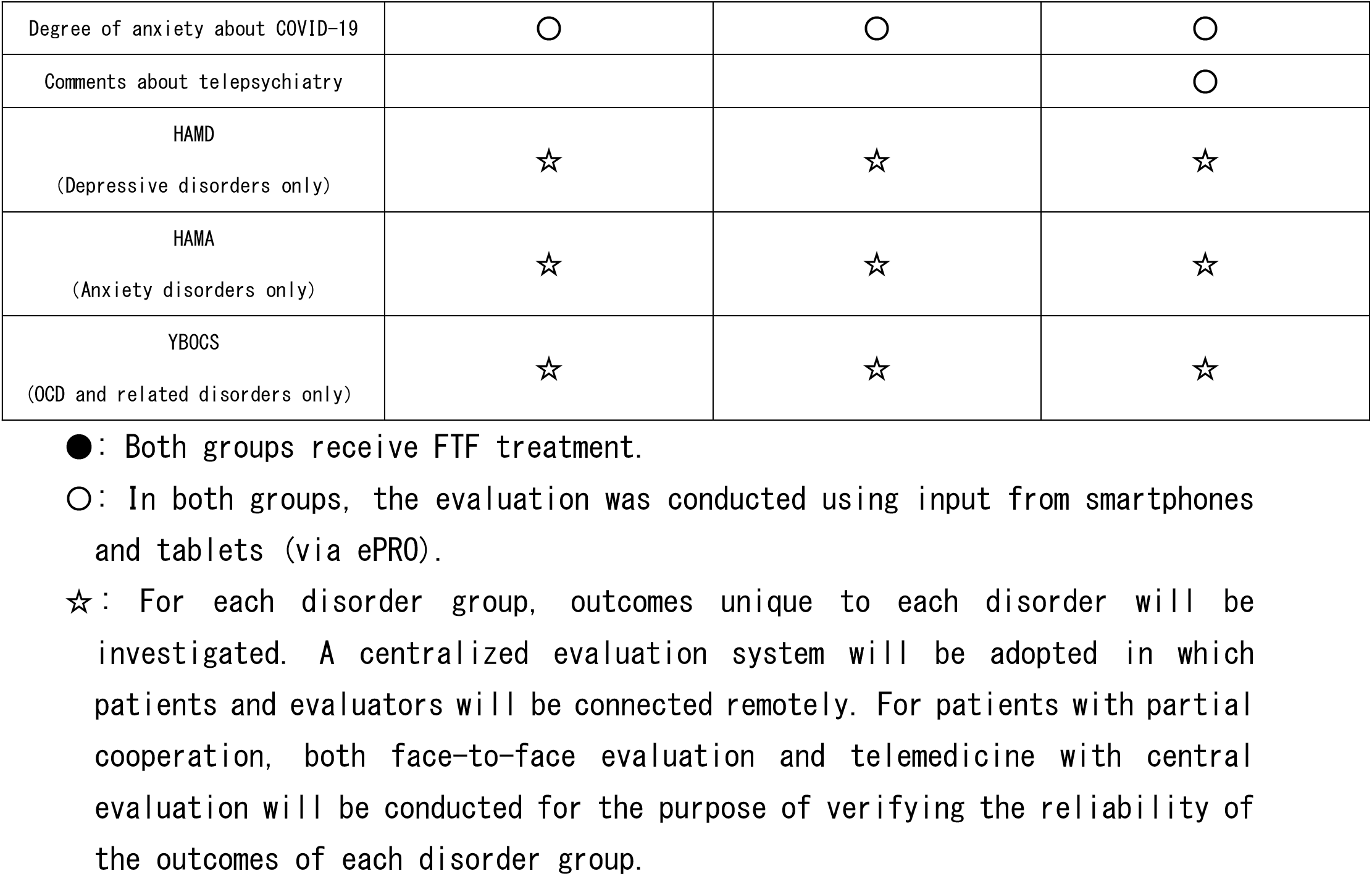
Schedule for data collection and evaluations during the study’s observation period.

### 2.5. Primary Outcomes

The primary outcome is the 36-Item Short Form Health Survey Mental Component Summary (SF36MCS) measured at baseline and at weeks 12 and 24. We will examine whether these changes differ between the Telepsychiatry and FTF groups. The SF36MCS is a self-administered rating scale to which patients respond via a dedicated application.

### 2.6. Secondary Outcome

The secondary outcomes are the following 12 items: 1) MOS 36-Item Short-Form Health Survey (SF36) (assessed at baseline and at weeks 12 and 24); 2) dropout rate (in the Telepsychiatry group, if the patient stops telepsychiatry and switches to FTF only, the patient will be considered to have dropped out of the Telepsychiatry group; 3) Working Alliance Inventory (WAI) scores as treatment alliance; 4) Client Satisfaction Questionnaire (CSQ) scores as satisfaction; 5) adverse events; 6) costs (Self-administered questionnaire on costs associated with medical visits, etc.); 7) EQ-5D (EuroQol 5 Dimension); 8) degree of anxiety about COVID-19; 9) comments about telepsychiatry (or central evaluation by telemedicine); 10) for the depressive disorder group, Hamilton Depression Rating Scale (HAMD); 11) for the anxiety disorder group, the Hamilton Anxiety Rating Scale (HAMA); and 12) for the OCD and related disorders group, the Yale-Brown Obsessive Compulsive Scale (YBOCS).

### 2.7. Sample Size

The primary outcome of the study is health-related quality of life as assessed using the SF36MCS, which will be evaluated at baseline and at weeks 12 and 24. The sample size is based on previous psychiatric intervention studies (including psychotherapy and electroconvulsive therapy interventions, but not studies comparing telepsychiatry and FTF), in which the evaluation period was 6 months [11–16]. In existing studies, the mean SF36MCS ranged from 30 to 50 and the standard deviation from 9 to 14.

In the present study, assuming that the SF36MCS of the telemedicine and FTF groups at 6 months is 45 (no difference between the two groups), with a standard deviation of 12, and a non-inferiority margin of 5, the required number of patients in each group would be 92 under the conditions of 80% power and a one-sided significance level of 2.5%.

In this study, the dropout rate is expected to be low because the primary psychiatrist who have been treating the patients up to the time of the study will continue to be in charge of the treatment regardless of whether the patients are treated in the Telepsychiatry group or in the FTF group. Assuming a dropout rate of about 10%, the total number of patients will be 200, or 100 in each group.

### 2.8. Data Collection and Management

Data on the SF36MCS, SF36, treatment alliance, satisfaction, cost, EQ-5D, and degree of anxiety about COVID-19 will be collected as self-administered patient-reported outcome measures. We will construct and operate a system to store these data directly using electronic data capture (EDC) and an electronic patient-reported outcome (ePRO). We will use “cubeCDMS” by CRScube Inc. for EDC and “cubePro” by CRScube Inc. for ePRO. The participants will install the ePRO application on their smartphones, etc., access the questionnaire included in the application, and enter and transmit the data. For the HAMD, HAMA, and YBOCS, remote centralized ratings performed through a video camera feed will be used. Evaluators must have completed a total of at least 30 hours of training on these evaluation items, and the evaluator will assess the examinees through a video camera feed at the time of each visit. In addition, as an optional part of the study, both FTF evaluations and centralized ratings for HAMD, HAMA, and YBOCS will be performed for patients who are willing to participate, so as to validate the centralized ratings.

### 2.9. Statistical analysis

The analyses of the primary and secondary outcomes will be performed in full analysis set (FAS), which includes all patients who took at least one SF36MCS assessment during the study, do not present any serious violation of the study protocol, and have data collected after treatment commencement. For the baseline characteristics, summary statistics will comprise frequencies and proportions for categorical variables, and means and standard deviations for continuous variables. The patient characteristics will be compared using a chi-square test for categorical variables, and a t test or Wilcoxon rank sum test for continuous variables.

For the primary analysis, aimed at comparing treatment effects, the least square mean and its 95% confidence interval (CI) will be estimated using a mixed-effects model for repeated measurements (MMRM). No imputation of missing data was performed. The MMRM model will include treatment, visit, treatment-by-visit interaction, and baseline SF36MCS. An unstructured covariance matrix will be assumed to model the within-patient variance and estimation was performed by restricted maximum likelihood method. Based on the model, the result of the SF36MCS at 6 months is expressed with point estimate for group difference and one-sided confidence interval with significance level of 2.5%. Non-inferiority is to be demonstrated if the upper boundary of this confidence interval does not exceed the non-inferiority margin of 5.

The secondary analysis will be performed in the same manner as the primary analysis. All comparisons are planned and all p values will be two sided. P values < 0.05 will be considered statistically significant. All statistical analyses will be performed using the SAS software version 9.4 (SAS Institute, Cary, NC, USA). The statistical analysis plan (SAP) will be developed by the principal investigator and the biostatistician before completion of patient recruitment and data fixation.

## 3. Results

The trial began in April 2021. Recruitment goals are on target to date, and the trial is projected to be completed in March 2023. The trial is registered at jRCT1030210037.

## 4. Discussion

The J-PROTECT study is an important study in both the psychiatric and telemedicine fields, as the COVID-19 outbreak is far from under control. The wide practice of telemedicine in this field is meaningful as an effective countermeasure against infection and to prove that telepsychiatry is effective for the treatment of psychiatric disorders that can worsen during a pandemic. As mentioned above, many studies comparing telepsychiatry with FTF treatment have focused on specific disorders or specific treatments, such as CBT, but few studies have examined multiple disorders simultaneously. For example, O’Reilly et al. randomly assigned 495 outpatients to FTF treatment or telepsychiatry and followed them for up to 4 months; they reported that telepsychiatry provided the same clinical outcome and satisfaction as FTF treatment, but at a lower cost [17]. In another study, De Las Curevas et al. randomly assigned 140 outpatients to FTF treatment or telepsychiatry with a 24-week follow-up period and reported that the efficacy of telepsychiatry was comparable to that of FTF treatment [18]. The novelty of the present study is that it will be the first pragmatic trial of telepsychiatry in Japan, a country that provides universal health insurance that allows free access to medical services with relatively low cost. It will also be a multi-center, multi-regional study, unlike previous pragmatic trials of multiple psychiatric disorders. Since the degree and content of telepsychiatry regulation varies in each country [6], showing that telepsychiatry is equally effective in different regulatory and cultural settings will be important to promote its use appropriately.

In considering the study design, several issues needed to be taken into account. First, the rationale for selecting SF36MCS as the primary outcome in this study was that it was necessary to establish an outcome that encompassed depressive disorder, anxiety disorder, and OCD and related disorders and that was in use at many psychiatric clinical studies as a health-related quality of life (QOL), which is a superordinate concept for the severity of all illnesses [11–16]. Next, the reason why we set the percentage of telemedicine in the telepsychiatry group to be more than 50% is to allow room for the use of both FTF and telepsychiatry depending on the patient’s condition, just as in regular medical care. The Japanese government guidelines also do not recommend the continuation of long-term treatment with telemedicine alone[19].

The limitations of this study are as follows. First, we targeted three disorder groups, namely depressive disorders, anxiety disorders, and OCD and related disorders. In addition to targeting three disorders in one trial, the demographic characteristics of the participants may vary over a wide range. This may lead to difficulty in finding a comparative effect size of telemedicine; however, this is an inherent difficulty of pragmatic trials, and limiting participation to a specific patient population may limit the generalizability of the results. Related to this point, IT literacy might be the largest modulator of the results, and since Japan is a world-leading aging society, this fact is of particular consideration. Elderly people generally experience more hurdles to telemedicine because of IT literacy issues [20]. By randomizing the whole population, we reduce the risk of the impact of this possible modulator. At the same time, by asking the participants regarding their impressions or hurdles regarding telemedicine utilization in a free-answer format, we hope to gather information that will enable us to help elderly or low-IT literacy patients to take advantage of telemedicine in the future. Finally, the implementation of this study may be affected by the status of the COVID-19 pandemic. If the pandemic becomes serious, there is a risk that hospitals will suspend FTF treatment or that patients will refuse to come to the hospital for fear of the risk of infection. In any case, we will implement an appropriate combination of telepsychiatry and FTF treatment, while prioritizing the interests of the patients.

## Data Availability

Due to the nature of this research, participants of this study did not agree for their data to be shared publicly, so supporting data is not available.

## Ethics approval and consent to participate

This study was approved by the Institutional Review Board of the National Center of Neurology and Psychiatry and the participating medical facilities. Researchers will obtain written informed consent from all the participants. Participants are able to leave the study at any time.

## Funding

This research is supported by the Japan Agency for Medical Research and Development (AMED) under Grant Number JP21dk0307098. The funding source did not participate in the design of this study and will not have any hand in the study’s execution, analyses, or submission of results.

Japan Agency for Medical Research and Development (AMED) 20F Yomiuri Shimbun Bldg. 1-7-1 Otemachi, Chiyoda-ku, Tokyo 100-0004 Japan Tel: +81-3-6870-2200, Fax: +81-3-6870-2241.

## Authors’ contributions

**Taishiro Kishimoto**:Conceptualization, Methodology, Writing - Review & Editing, Funding acquisition, **Shotaro Kinoshita**: Writing - Original draft preparation. **Shogyoku Bun**: Methodology, Investigation, Writing - Review & Editing, **Yasunori Sato**: Methodology, Formal analysis, **Momoko Kitazawa**: Methodology, Writing - Review & Editing, **Toshiaki Kikuchi**: Methodology, Investigation, **Mitsuhiro Sado**: Methodology, Investigation, **Akihiro Takamiya**: Methodology, Investigation, **Masaru Mimura**: Supervision, **Takashi Nakamae**: Methodology, Investigation, **Yoshinari Abe**: Methodology, Investigation, **Tetsufumi Kanazawa**: Methodology, Investigation, **Hiroaki Tomita**: Methodology, Investigation, **Koichi Abe**: Methodology, Investigation, **Akitoyo Hishimoto**: Methodology, Investigation, **Takeshi Asami**: Methodology, Investigation, **Akira Suda**: Methodology, Investigation, **Yoshinori Watanabe**: Methodology, Investigation, **Toru Amagai**: Methodology, Investigation, **Kei Sakuma**: Methodology, Investigation, **Hisashi Kida**: Methodology, Investigation, **Michitaka Funayama**: Methodology, Investigation, **Hiroshi Kimura**: Methodology, Investigation, **Aiko Sato**: Methodology, Investigation, **Soichiro Fujiwara**: Methodology, Investigation, **Kiichiro Nagao**: Methodology, Investigation, **Naoya Sugiyama**: Methodology, Investigation, **Maki Takamiya**: Methodology, Investigation, **Hideyuki Kodama**: Methodology, Investigation, **Takaharu Azekawa**: Methodology, Investigation.

All authors critically revised the manuscript, and approved the manuscript to be published, and agree to be accountable for all aspects of the work in ensuring the questions related to the accuracy or integrity of any part of the work are appropriately investigated and resolved.

## Declaration of competing interest

The authors declare the following financial interests/personal relationships which may be considered as potential competing interests:

T. Kishimoto: Research grant funding, Micin.

T. Nakamae: Provided with a telemedicine system, Medley.

Y. Abe: Provided with a telemedicine system, Medley.

## Acknowledgements

The authors are grateful to Dr. Kazunari Yoshida, Dr. Toshiro Horigome, Dr. Mayu Fujikawa, Ms. Kumiko Hiza, Ms. Hiromi Mikami, Ms. Saki Hattori, Mr. Masao Takaishi, Mr. Satoshi Tsujimura, Mr. Hajime Tamura, Ms. Junko Suzuki, Ms. Keiko Komiyada, Dr. Anri Watanabe, Dr. Yoshihiro Matsumoto, Ms. Satoko Kimura, Ms. Haruka Okamoto, Dr. Kyosuke Sawada, Ms. Yuka Oba, Mr. Satoshi Tsujimura, Ms. Shii Sagae, Mr. Kiyoji Nagao, Mr. Ryuhei Terashi, Ms. Sumako Onishi, Ms. Mayumi Hiruma, Ms. Junko Kannnari, Ms. Kanako Sasao, Ms. Ayumi Konishi, Ms. Nobuko Haga, Mr. Nobuhiko Noguchi, Mr. Kosuke Hino, Mr. Yuya Igarashi, Dr. Kenichi Goto for supporting data collection and management in this study.

## Abbreviations

COVID-19: the 2019 novel coronavirus disease
J-PROTECT: Japanese Project for Telepsychiatry Evaluation during COVID-19: Treatment Comparison Trial
DSM-5: Diagnostic and Statistical Manual of Mental Disorders, Fifth Edition
FTF: face-to-face treatment
jRCT: Japan Registry of Clinical Trials
OCD: obsessive–compulsive disorder
SF36MCS: MOS36-Item Short-Form Health Survey Mental Component Summary
SF36: MOS 36-Item Short-Form Health Survey
WAI: Working Alliance Inventory
CSQ: Client Satisfaction Questionnaire
EQ-5D: EuroQol 5 Dimension
HAMD: Hamilton Depression Rating Scale
HAMA: Hamilton Anxiety Rating Scale
YBOCS: Yale-Brown Obsessive Compulsive Scale
EDC: electronic data capture
ePRO: electronic patient-reported outcome
QOL: quality of life

## Notes

### Clinical Trial

jRCT1030210037

### Author Declarations

This study was approved by the Institutional Review Board of the National Center of Neurology and Psychiatry and the participating medical facilities.

